# Covid-19 impact on Parkinson’s Disease patients treated by drugs or deep brain stimulation

**DOI:** 10.1101/2021.03.03.21252464

**Authors:** Mehri Salari, Masoud Etemadifar, Alireza Zali, Zahra Aminzade, Parsa Farsinejad, Sepand Tehrani Fateh

**Affiliations:** Functional Neurosurgery Research Center, Shohada Tajrish Comprehensive Neurosurgical Center of Excellence, Shahid Beheshti University of Medical Sciences, Tehran, Iran; Department of Functional Neurosurgery, Medical School, Isfahan University of Medical Sciences, Isfahan, Iran; Chancellery, Shahid Beheshti University of Medical Sciences, Tehran, Iran; School of Medicine, Shahid Beheshti University of Medical Sciences, Tehran, Iran

**Author notes:** Corresponding Author: Sepand Tehrani Fateh, Address: Koodakyar st., Daneshjoo Blvd., Velenjak, Tehran, Iran.

**Keywords:** Parkinson’s Disease, Deep Brain Stimulation, Covid-19, SARS-CoV-19, Pandemic

## Abstract

**Purpose:** Covid-19 has affected all people, especially those with chronic diseases, including Parkinson’s Disease (PD). Covid-19 may affect both motor and neuropsychiatric symptoms of PD patients. We intend to evaluate different aspects of Covid-19 impact on PD patients.

**Methods:** 647 PD patients were evaluated in terms of PD-related and Covid-19-related clinical presentations in addition to past medical history during the pandemic through an online questioner. They were compared with an age-matched control group consist of 673 individuals and a sample of the normal population consist of 1215 individuals.

**Results:** The prevalence of Covid-19 in PD patients was 11.28%. The mortality was 1.23% among PD patients. The prevalence of Covid-19 in PD patients who undergone DBS was 18.18%. No significant association was found between the duration of disease and the prevalence of Covid-19. A statistically significant higher prevalence of Covid-19 in PD patients who had direct contact with SARS-CoV-19 infected individuals was found. No statistically significant association has been found between the worsening of motor symptoms and Covid-19. PD patients and the normal population may differ in the prevalence of some psychological disorders, including anxiety and sleeping disorders, and Covid-19 may affect the psychological status.

**Conclusion:** PD patients possibly follow tighter preventive protocols, which lead to lower prevalence and severity of Covid-19 and its consequences in these patients. Although it seems Covid-19 does not affect motor and psychological aspects of PD as much as it was expected, more accurate evaluations are suggested in order to clarify such effects.

## Introduction

In December 2019, the severe acute respiratory syndrome coronavirus 2 (SARS-CoV-2) or Covid-19, causing a pneumonia-like illness, emerged in China and spread quickly all around the world and now is an ongoing pandemic [1]. The effect of the neurological disorder as a risk factor for Covid-19 infection is yet to be fully understood, and its impact on clinical features of these diseases need to be well characterized [2].

Parkinson’s disease (PD) is one of the most common neurodegenerative disorders of the central nervous system, with an increased prevalence by aging [3]. As the PD prevalence is related to the age and elderly and those with comorbidities are more vulnerable to Covid-19 infection, it can be suggested that Covid-19 infection risk might be increased in PD patients [4]. Moreover, Covid-19 can also affect motor and neuropsychiatric symptoms of PD patients [5, 6].

In this study, we evaluated the prevalence of Covid-19 in 647 PD patients along with other parameters, including mortality during the pandemic, Covid-19 symptoms, alteration in PD motor and psychological symptoms.

## Methods

### Study Design and Population

This cross-sectional, case-control study was approved by the Iran National Committee for Ethics in Biomedical Researches (IR.SBMU.RETECH.REC.1398.439) to investigate Covid-19 prevalence and death rate its clinical features, and further consequences in Parkinson’s disease patients compared to the general population. The studied population was 647 patients diagnosed with PD based on UK Brain Bank criteria who were being followed-up at the Referral Movement Disorder Center in Shohada Tajrish hospital, Tehran. The study carried out in November 2020, when Iran was in the third pick of the coronavirus pandemic. After obtaining informed consent from all patients, a questionnaire which was prepared for the patients, consists of demographic data, medical history, and detailed PD-related and Covid-19-related clinical information of the patients, was completed over the phone call. The same questionnaire was designed for the control groups and our sample of normal population which has been broadcasted over the internet and social media. The questionnaire was answered by 1366 individuals, and among those, 673, and 1215 individuals selected as the age-matched control group and a sample of the normal population, respectively. No identifiable data was not included in the questioner such as name, phone number, national ID numbers, etc., for the control group.

### Statistical analysis

Statistical analysis tests were as follows: Fisher Exact Text (for categorical variable), Mann-Whitney U test (for continuous variables), and Anderson-Darling test (assessment of distribution). Statistical analysis was done with Excel Microsoft office, and R language (4.0.2 ed,2020) implemented in R studio (1.3.1093 ed,2020). Significance for all statistical analyses was fixed at 0.05.

## Results

### Mortality and Covid-19 prevalence

In this study, we evaluated 647 PD patients (Mean age: 60.57±12.46, Male: 410, Female: 237) for Covid-19 prevalence of their Covid-19- and PD-related presentations. 1215 individuals (Mean age: 39.84±14.90) were considered as a sample of the normal population, and 673 individuals (Mean age: 56.32±11.62) were selected as an age-matched control group. The prevalence of Covid-19 among 647 PD patients was 11.28%, and it was significantly lower (p-value<0.05) than the normal population (15.39%). However, the difference of Covid-19 prevalence between PD patients and the control group (14.85%) was statistically insignificant (p-value≈ 0.0605). Mortality of PD patients was 1.23% (stroke: 1, cardiac arrest without definite diagnosis: 2, car accident: 1, gastrointestinal bleeding: 1, Unknown: 3) (Table1). Ten PD patients (13.69%) were hospitalized because of the severity of Covid-19, while this rate was about 5.34% for the normal population and the difference between these hospitalization rates was statistically significant (P-value<0.05). On the other hand, eighteen individuals (18%) in the control group were hospitalized because of the severity of Covid-19, and the hospitalization rate of this group was not significantly different from that of PD patients (p-value>0.5). 44 of 647 PD patients were undergone DBS surgery, and 8 of them (18.18%) got infected with SARS-CoV-19; there was no significant difference between the prevalence of Covid-19 in these patients with either the rest of PD patients or the normal population.

**Table1.**

| **Table1** |  |  |  |  |  |
| --- | --- | --- | --- | --- | --- |
| **Demographics and Covid-19 Prevalence** | **Mean age±SD** | **Median age (min, max)** | **Mean Duration of PD** | **Male:Female** | **Covid-19 Prevalence** |
| **PD** | 60.57±12.46 | 62(17, 92) | 7.67±6.04 | 410:237 | 11.28% |
| **DBS** | 56.13±8.72 | 57(34, 70) | 14.52±6.68 | 25:19 | 18.18% |
| **Normal** | 39.84±14.90 | 36(3, 93) | N/A | 455:760 | 15.39% |
| **Control** | 56.32±11.62 | 55(40, 93) | N/A | 280:393 | 14.85% |
| **Underlying diseases (2 most frequent)** |  |  |  |  |  |
| **PD** | hypertension (25.19%) and diabetes (17.15%) |  |  |  |  |
| **Control** | hypertension (26.44%) and diabetes (14.26%) |  |  |  |  |

Demographic information, Covid-19 prevalence and two most frequent underlying diseases of PD and control group. (N/A: Not applicable)

**Table2.**

| **Table2** |  |  |  |  |  |  |
| --- | --- | --- | --- | --- | --- | --- |
| **#** | **Sex** | **Age** | **Duration of PD** | **Underlying disease** | **Family History** | **Cause of death** |
| **1** | Male | >70 | 10-15 | DM | Positive | Unknown |
| **2** | Male | >70 | ≤10 | Seizure | Negative | Stroke |
| **3** | Male | 60-70 | ≥15 | BPH, DM | Negative | Unknown |
| **4** | Female | >70 | ≤10 | - | Negative | Accident |
| **5** | Female | 60-70 | ≥15 | Hypertension | Negative | GI Bleeding |
| **6** | Male | 60-70 | 10-15 | DM, Depression | Negative | Cardiac arrest |
| **7** | Female | <60 | ≤10 | - | Negative | Unknown |
| **8** | Female | <60 | ≥15 | - | Negative | Cardiac arrest |

Mortality cases among PD group. (BPH: Benign Prostatic Hyperplasia, DM: Diabetes Mellitus, GI: Gastrointestinal)

### Past medical history of PD patients

Tremor, bradykinesia, and rigidity were the most frequent primary symptoms of PD patients, respectively, in our study. 272 PD patients had a family history of neurological disorders which the four most frequent of them was a family history of PD in 156 patients, a family history of Alzheimer Disease (AD) in 71 patients, a family history of migraine in 64, and a family history of Multiple Sclerosis (MS) in 34. No significant association was found between having a family history of PD, MS, and migraine and higher/lower prevalence of Covid-19; However, interestingly, it has been found that such an association exists between having the family history of Alzheimer and a higher prevalence of Covid-19 (P-value<0.05). 32 of 647 PD patients got influenza vaccine during the last season. No significant difference has been found in the Covid-19 prevalence, neither between PD patients who got influenza vaccine and PD who did not (P-value>0.7) nor between PD patients and normal population who got influenza vaccine (P-value>0.4). Moreover, there was no significant difference between the prevalence of Covid-19 in PD patients and the control group who both got influenza vaccine. It worth noting that the normal population and the control group were vaccinated for influenza more than PD patients (P-value<0.05 and P-value<0.01 respectively). Duration of PD was about as following: (<1year: 9.4%, 1-5years: 37.09%, 5-10years: 29.05%, >10years: 24.11%) and we could not find any statistically significant association between duration of disease and prevalence of Covid-19 (P-value>0.05).

### Covid-19 history

Myalgia (45.30%), fever (41.09%), fatigue and weakness (39.72%), cough (35.61%), headache (23.28%), dyspnea (21.91%), Congestion or runny nose (17.80%), sore throat (13.69%) and anosmia (12.32%) were most frequent symptoms of Covid-19 in the PD patients, respectively while they were fever (55.08%), fatigue and weakness (55.08%), myalgia (54.01%), cough (45.98%), headache (43.85%), anosmia (42.78%), loss of appetite (35.82%) and sore throat (35.29%) in the normal population respectively. PD patients reported significantly lower occurrence of viral infections before the Covid-19 pandemic in comparison to the normal population (P-value<0.05), but there was no significant difference between the occurrence of viral infections before the Covid-19 pandemic in PD patients and the control group (P-value>0.1). We found a statistically significant higher prevalence of Covid-19 in PD patients who had direct contact with SARS-CoV-19 infected individuals (72.06%) and that of the normal population with a similar condition (P-value<0.05). However, there was not a significant difference between the prevalence of Covid-19 in PD patients who had direct contact with SARS-CoV-19 infected individuals and that of the control group with a similar condition (63%) (p-value>0.1). During the pandemic, 71.87% of PD patients visited their physicians at hospitals or offices.

### PD motor symptoms

32.92% of PD patients have reported worsening of motor symptoms, while this rate was 38.65% in PD patients who had undergone a DBS surgery and 41.09% in PD patients who were infected with Covid-19. No statistically significant difference has been found in worsening of motor symptoms between PD patients who had Covid-19 and PD patients who did not (P-value>0.1). Moreover, no statistically significant difference has been found in worsening of motor symptoms between PD patients who had undergone a DBS surgery and PD patients who had not (P-value>0.4). Also, no significant association between the rate worsening of motor symptoms and the rate of visiting physicians was observed (P-value>0.9).

### Psychological evaluations

43% and 41.09% of all PD patients and PD patients who had Covid-19, respectively, reported deterioration in their psychological status such as occurrence or worsening of depression. No statistically significant difference has been found in the rate of deterioration in psychological status between PD patients who had and had not Covid-19 (P-value>0.7). Such difference was also statistically insignificant between Covid-19-infected PD patients and either Covid-19-infected normal population or Covid-19-infected control group (P-value>0.4 and P-value=1 respectively). There was a statistically significant difference between the rate of deterioration in psychological status between all PD patients and the normal population and also all PD patients and the control group (P-value<0.01 and P-value<0.01 respectively), and this rate were higher in the PD patients in both comparisons. Moreover, this rate was significantly higher in PD patients who were not infected with Covid-19 in comparison to this rate in not infected individuals in the normal population (P-value<0.01).

46.21% of all PD patients and 52.05% of PD patients who had Covid-19 declared that they had experienced anxiety during the pandemic. The prevalence of anxiety was significantly lower in PD patients in comparison to the normal population (P-value<0.05). Moreover, a statistically significant difference was present between the rate of anxiety in all PD patients and in the control group (P-value<0.05). No significant difference was observed in the prevalence of anxiety between PD patients who had Covid-19 and PD patients who had not Covid-19 (P-value>0.05). Moreover, the prevalence of anxiety in PD patients who had Covid-19 and the normal population who had Covid-19 was not significantly different (P-value>0.5). On the other hand, the difference between the prevalence of anxiety in PD patients who had Covid-19 and the control group who had Covid-19 was not statistically significant (P-value>0.5). In addition, no association was observed between the hospitalization of PD patients subsequent to Covid-19 and the prevalence of anxiety in these patients (P-value>0.5).

18.54% of all PD patients and 26.02% of PD patients who had Covid-19 reported sleeping problems during the pandemic. Both the normal population and Covid-19-infected portion of the normal population demonstrated significantly higher rates of sleeping problems in comparison to all PD patients and PD patients who had Covid-19, respectively (P-value<0.01). However, there was no statistically significant difference between the rate of sleeping problems in PD patients and the control group (P-value>0.08), and also, there was no such difference between PD patients who had Covid-19 and the control group who had Covid-19 (P-value>0.1). However, the rate of sleeping problems had no significant association with neither rate of hospitalization nor prevalence of Covid-19 (P-value>0.1).

## Discussion

The occurrence of Covid-19 and its complications in patients with underlying diseases were among the very first concerns. The likelihood of hospitalization and death in patients with movement disorders after Covid-19 is found to be greater in comparison with the general population.

Different reports have been published concerning the prevalence of Covid-19 in PD patients; however, consistency between results cannot be seen in some cases. A study showed that Covid-19 prevalence in PD patients was similar to the controls, and the difference in mortality rates between the PD population and the control were not significant. However, considering the methodology of this study, it can be said that some patients might be passed away or hospitalized in other facilities and hence unreachable for monitoring and data-collecting procedures [6]; therefore, the mortality rate might be quite inaccurate. In this study, the PD patients infected with Covid-19 were younger, more frequently obese, and suffering from chronic obstructive pulmonary disease in comparison to not infected patients. The younger age of this group may be due to the lower preventive measures of young people [7].

In contrast, in another study, the expected rate of deaths in hospitalized patients with idiopathic PD found to be higher than that of PD patients before the pandemic period [8], suggesting that Covid-19 may play a role. Delay in seeking medical attention is suggested as a possible cause for such an outcome [8]. It is also reported that PD patients were at a significantly higher risk of dying from Covid-19 compared to patients without PD [9]. Tuscany study on 740 PD patients revealed Covid-19 prevalence of 0.9% among these patients in addition to 0.13% mortality. The prevalence of Covid-19 was higher in respect to that of in general population in Tuscany and Italy [9]. Hypertension and diabetes were more prominent in PD patients affected with Covid-19 compare to non-Covid-19, indicating these two comorbidities may act as risk factors for Covid-19 in PD [10]. Moreover, either motor or non-motor symptoms did not worsen in PD patients during the lockdown period [11]. During a large cohort (1407 patients) conducted in Piedmont-Italy, the prevalence of Covid-19 in interviewed PD patients was found to be 0.57%, while Covid-19 prevalence was 0.63% for the general population in the same region. It is reported that 75% of PD patients lost their lives because of Covid-19 in comparison to the mortality rate of 11.53% in the general population [11]. The similarity between PD patients and the general population’s Covid-19 prevalence may be due to higher levels of attention, percussion and self-isolation among these patients [11]. It is suggested that older age and longer disease duration might be predictors of worse outcomes as the mortality rate is higher among older patients with a longer duration of disease [12]. In the last study, PD symptoms worsened in all patients, and fever (100%), weakness (43%), muscle pain (29%), dyspnea (29%), and cough (14%) were the most frequent PD symptoms, which got worsen early before or early after the onset of Covid-19 signs [12]. Wuhan’s study reported a 1.77% prevalence of Covid-19. Moreover, 3 out of 509 (0.58%) PD patients diagnosed with Covid-19 died. Old age, chronic comorbidities, neurological complications, and sudden discontinuation of anti-parkinsonian drugs were recognized as potential risk factors for mortality in Covid-19 patients with PD [12]. Another study reported a 2.6% prevalence of Covid-19 in PD patients with zero mortality. It is reported that more than half of the patients experienced worsening of the symptoms, which were as the following: bradykinesia 47.7%, sleep problems 41.4%, rigidity 40.7%, gait disturbances 34.5%, anxiety 31.3%, pain 28.5%, fatigue 28.3%, depression 27.6%, tremor 20.8%, and appetite disorders 13.2% [12]. Cardiovascular diseases are suggested as possible risk factors in worse outcomes in PD patients affected with Covid-19 [13]. A different mortality rate is reported in another study in which a multi-center cohort revealed 19.7% overall mortality among PD patients diagnosed with Covid-19. Dementia, hypertension, and PD duration may be potential predictors for such outcomes [14].

Most studies have an agreement on the possibly higher Covid-19 prevalence and Covid-19-related mortality in PD patients, while some others have predicted such outcome considering the pathophysiological processes of both Covid-19 infection and PD. It has been suggested that susceptibility of PD to Covid-19 may be related to either the pathophysiology of PD, factors that are subsequent to the current condition of the health system during the pandemic, or direct and indirect health effects of Covid-19 on PD patients. In our study, the prevalence of Covid-19 among PD patients was 11.28% which was significantly lower than that of the normal population. A lower prevalence of Covid-19 in PD patients was not expected as it has been hypothesized in many articles that PD patients are at higher risk for being infected with SARS-CoV-19 due to some reasons as below. Pathophysiological processes of PD lead to respiratory complications [14], gastrointestinal problems [15], sedentary lifestyle [4, 16], immune systems malfunction [16], nervous systems dysfunction [4, 16], and a need for using medications with possible side effects and drug interactions [17], may place PD patients at higher risks of mortality and morbidity subsequent to Covid-19 infection. However, the effect of anti-PD drugs was found to be insignificant on the prevalence of Covid-19 [17]. In contrast, in another study, sudden discontinuation of anti-parkinsonian drugs is recognized as a potential cause of death in PD patients infected with Covid-19 [16, 18]. Advanced age of PD patients and long disease duration would be other risk factors for vulnerability to Covid-19 [8]. However, the lower prevalence of Covid-19 in PD patients in comparison to the normal population in our study may be due to more aggressive preventive protocols that these patients follow. Moreover, in line with the last statement, the observation that the prevalence of Covid-19 in PD patients and the control group was not significantly different, can be interpreted as a result of possible aggressive preventive protocols that the individuals in the control group would follow because of their underlying diseases or advanced ages. This suggestion can be strengthened by the result that also the prevalence of other viral infections in PD patients before the pandemic was lower than the normal population. Moreover, these results may not be interpreted as the consequences of the pathophysiology of PD or its medications, as we have observed that PD patients who had first degree relatives infected with Covid-19 possessed a higher prevalence of Covid-19 in comparison with the same group in the normal population. This result indicates that possibly PD patients are at higher risk of being infected with Covid-19. We also analyzed the prevalence of Covid-19 in a group of PD patients who had undergone a DBS surgery, and it seems that DBS surgery might not put PD patients at higher risk of Covid-19.

We found that although PD patients were hospitalized because of the severity of Covid-19 more than our normal population, the rate of hospitalization among these patients was not significantly different from our control group. This may indicate that other causes are involved in increasing the rate of hospitalization rather than PD. Our observation is in line with a paper from Vignatelli et al., which states that “PD per se probably is not a risk factor for Covid-19 hospitalization” [19].

The current condition also imposes limitations which may lead to deterioration in PD clinical course. Reduction in physical activity during quarantine may lead to worsening of both motor and non-motor symptoms as exercise may attenuate clinical symptom progression in PD [13]. Availability of PD medications, health services, and physicians’ advice or guidance during the lockdowns is recognized as an important factor in the deterioration of the clinical course of PD patients. In this sense, disability in PD patients may increase due to a lack of access to suitable and sufficient medications. This condition may lead to an increase in motor, non-motor, and neuropsychiatric symptoms and possibly higher mortality [17, 18].

In our study, 32.92%, 38.65%, and 41.09% of PD patients, PD patients who had undergone a DBS surgery, and PD patients who were infected with Covid-19 reported worsening of motor symptoms. However, we could not find any significant between Covid-19 infection and worsening of motor symptoms or between DBS surgery and worsening of motor symptoms. Moreover, we could not find any association between a deterioration in psychological status and Covid-19 in PD patients. Such association could not also be found between anxiety and Covid-19 and sleeping disorders and Covid-19. But interestingly, the normal population demonstrated higher levels of deterioration in psychological status, anxiety, and sleeping disorders than PD patients. Since our normal population was significantly younger than the population of PD patients, it is possible to interpret these results as a reflection of higher levels of psychological tensions that younger people encounter with. This interpretation can be strengthened with the observation that PD patients had significantly higher levels of deterioration in psychological status and anxiety than the control group with a similar mean age. Therefore, it can be said the PD patients may have higher levels of deterioration in psychological status and anxiety than the control group due to some unknown underlying mechanisms in the pathophysiology of PD, but these two are not as high as they are in the normal population because of possibly more psychological tensions.

Since some PD patients could not be reached because they may have passed away or hospitalized in other facilities, the prevalence of Covid-19 and the rate of mortality might be slightly different in comparison with the condition that these points are taken into account. Another limitation of our study would be the inaccuracy of our control group in terms of gender and age distribution in spite of similar mean age.

## Conclusion

Patients with chronic diseases including, PD, are thought to be at higher risks of infection and its complications during the Covid-19 pandemic. Moreover, Covid-19 may affect different aspects of PD, including motor and psychological signs and symptoms, but in a different way unexpectedly.

## Data Availability

All data would be available on a request.

## Acknowledgement

This article is taken from disease registry, titled “Parkinson’s Disease Registry in Patients Referred to Neurology Clinics of Shahid Beheshti University of Medical Sciences in Tehran (SBMU-PDR)” and code number “IR.SBMU.RETECH.REC.1398.439” from ethic committee that was supported by deputy of research and technology in Shahid Beheshti University of Medical Sciences (http://dregistry.sbmu.ac.ir).

## Authors’ Roles

Conceptualization and study design: MS

Data collecting: MS, ME, PF

Statistical Analysis: ST

Literature Review: ST, ZA

Writing Manuscript: ST, ZA

Editing: MS, ST

Supervision: MS, AZ, ST

## Notes

**Financial disclosure:** None of the authors have received any funding of any kind for either this project or other projects.

**Conflicts of interest:** Authors declare no conflicts of interest.

### Competing Interest Statement

The authors have declared no competing interest.

### Funding Statement

None of the authors have received any funding of any kind for either this project or other projects.

### Author Declarations

This cross sectional, case-control study approved by the Iran National Committee for Ethics in Biomedical Researches (IR.SBMU.RETECH.REC.1398.439)

